# Building Medical Rehabilitation System with Safety and Without Interruption for the Patients with Coronavirus Disease 2019 Using the Functional Resonance Analysis Method (FRAM)

**DOI:** 10.1101/2022.05.18.22275242

**Authors:** Naoki Sasanuma, Keiko Takahashi, Ai Yanagida, Yohei Miyagi, Seiya Yamakawa, Tetsu Seo, Yuki Uchiyama, Norihiko Kodama, Kazuhisa Domen

**Affiliations:** Department of Patient Safety and Quality Management, Hyogo Medical University, Nishinomiya, Japan; Department of Rehabilitation, Hyogo Medical University Hospital, Nishinomiya, Japan; Department of Rehabilitation Medicine, Hyogo Medical University, Nishinomiya, Japan

**Keywords:** Functional Resonance Analysis Method, FRAM, COVID-19, rehabilitation medicine

## Abstract

**Introduction:** Coronavirus disease 2019 (COVID-19) is an indication for rehabilitation medicine, especially in severe cases. However, there has been no systematic development of a safe and uninterrupted provision system of medical rehabilitation for patients and medical staff with COVID-19. The Functional Resonance Analysis method (FRAM) is used to analyze performance in a socio-technical system. In FRAM, each “Function” is viewed from six aspects : Input, Output, Preconditions, Resources, Control, and Time. These aspects define each Function and reveal connections between Functions. In this study, we analyzed a safe and uninterrupted provision system for medical rehabilitation—for severely ill COVID-19 patients using FRAM to prepare for possible problems in the future.

**Methods:** The subject of analysis was the provision system for medical rehabilitation for patients with COVID-19 at the Rehabilitation Center of Hyogo College of Medicine College Hospital. The analysis was conducted by dividing a 21-month rehabilitation period beginning April 2020 into 5 phases, and analyzing each phase using FRAM. The first four phases were retrospective analyses, and the fifth phase was a prospective analysis.

**Results:** Our results showed that the number of rehabilitation physicians, consultation systems, and full-time therapists was adjusted and the system providing rehabilitation was modified during each phase.

**Discussion:** Elements of Function, such as preconditions, control, and resources, require modification in each phase. In the process of adding and deleting these elements, it became clear that it was necessary to deal with new characteristics of SARS-CoV-2 infection. Retrospective system analysis using FRAM may contribute to the planning of measures necessary for the implementation of rehabilitation medicine prospectively.

## INTRODUCTION

The SARS-CoV-2 virus is highly transmissible and coronavirus disease 2019 (COVID-19) has affected a large number of people [1]. Because of the high transmissibility, medical institutions are required to treat COVID-19 cases separate from other patients, with medical staff wearing personnel protective equipment (PPE) to reduce the risk of infection. While more than 60% of COVID-19 cases are considered mild [2], some cases become very severe and require ventilatory management or extracorporeal membrane oxygenation. In these severe cases, lung injury induced by spontaneous breathing has been reported [3], and deep and/or rapid spontaneous breathing that requires effort may lead to progressive lung injury [3]. In cases with severe lung injury that requires ventilatory management, a sufficient amount of sedation is administered [4]. Suppression of spontaneous respiration for several days to several weeks during deep sedation prevents the exacerbation of pulmonary injury, however, the synergistic effects of immobility and sedation during this period cause the functional decline of systemic skeletal muscles in approximately 80% of patients [5]. This weakness of skeletal muscle has been reported previously as intensive care unit-acquired weakness [6], indicating the need for early physical therapy and rehabilitation care in the intensive care unit [7].

Our institution is a long-term acute care hospital that provides emergency and acute care in a region of Japan with a population of 1.7 million. Since March 2020, we have provided medical care by admitting only patients with COVID-19 who are severely ill or are likely to become severely ill. Since COVID-19 patients suffer from a decline in physical motor functions as described above, we have been providing medical rehabilitation since April 2020 [8]. Rehabilitation professionals, such as physical therapists, occupational therapists, and speech pathologists provide exercise training, assisted practice of basic activities, practice of activities of daily living, and, in some cases, unmasking of the patient and evaluation of various exercises in the oral and pharyngeal regions. Rehabilitation professionals in charge of COVID-19 cases wear full-PPE and instruct exercise training, but the combination of the virus’ ability to transmit infection and the density of contact between rehabilitation professionals and patients (a concept that combines contact time, distance, and amount of aerosol exposure) may put therapists at higher risk of infection [9]. Some reports have recommended specific PPE, but have not demonstrated its safety against the infectious potential of new emergent strains or safety in terms of therapist-patient distance and contact time [10, 11].

The Functional Resonance Analysis Method (FRAM) [12] is a systems analysis method developed by Hollnagel et al. and has been applied in medicine to analyze the occurrence of accidents and to solve clinical problems [13], including clinical problem solving under COVID-19 epidemic conditions [14]. In order to provide medical rehabilitation intervention uninterrupted and safely to patients with COVID-19, we have attempted to construct a system providing rehabilitation based on the number of patients, disease severity, infectivity of the SARS-CoV-2 virus, and health status of the staff. In this study, we used FRAM in the process of constructing the medical rehabilitation providing system, to understand its challenges, to clarify measures used to address challenges, and to predict future challenges and necessary measures.

The purpose of this study was to use FRAM to analyze a safe and continuous system to provide medical rehabilitation to patients with severe COVID-19, and to prepare for possible challenges that may arise in the future.

## METHODS

This study was approved by the Ethics Committee of Hyogo Medical College (registration number 3866) and was described in line with the SQIURE (Quality Improvement Reporting Excellence) 2.0 checklist[15].

This study was performed as a retrospective observational study. The subject of analysis was the provision system for medical rehabilitation to COVID-19 patients at the Rehabilitation Center of Hyogo Medical University Hospital. The analysis period was from April 2020 to December 2021. The Department of Rehabilitation consists of 5 rehabilitation physicians, 4 resident physicians, 36 physiotherapists, 14 occupational therapists, 7 speech pathologists, and 1 support staff. In the analysis process, a physician affiliated with the Department of Medical Safety and Quality Management provided guidance on the methods and validity of the analysis.

The analysis was conducted by dividing the target period into five phases:

Phase 1: FRAM construction of a system providing conventional medical rehabilitation before accepting COVID-19 cases,

Phase 2: From the beginning of COVID-19 case acceptance to the third month,

Phase 3: From the 4th month to the 8th month,

Phase 4: From the 9th month to the 21st month; FRAM analysis was conducted to examine the changes in the rehabilitation system in each of the phases and the problems and countermeasures in each period were confirmed.

Phase 5: Based on the FRAM analyses up to Phase 4, we attempted to examine the problems expected after the 22nd month and to plan countermeasures.

FRAM analysis was conducted by describing functions and analyzing diagrams according to the procedure described by Hollnagel [16]. The analysis procedure for each phase was as follows: 1) Export function names and each functional element and create FRAM diagrams (Phase 1); 2) Describe new functions that have arisen with the acceptance of COVID-19 cases and illustrate inter-functional linkage (Phase 2); 3) Describe the problems that occurred during system continuity and illustrate the linkage between functions (Phase 3-4); 4) Anticipate future system problems and responses (Phase 5).

In order to understand the characteristics of the patient population, the following items were recorded from the medical records: age, gender, height, body mass, body mass index (BMI), Quick COVID-19 Severity Index (qCSI)[17], hospitalization period, duration of medical rehabilitation, mechanical ventilation use, and high flow oxygen therapy use. Serum albumin, AST, ALT, LDH, CRP, white blood cell count, lymphocyte percentage, D-dimer, and ferritin were recorded from biochemical data at the time of admission to assess the severity of each patient’s illness [18–20].

## RESULTS

Between April 2020 to December 2021, 155 patients with COVID-19 received medical rehabilitation at the Rehabilitation Center of Hyogo College of Medicine College Hospital. A summary of the 115 cases is shown in Table 1. Of these cases, 29.6% received high-flow oxygen therapy and 36.5% received mechanical ventilation, indicating a population with a high proportion of severely ill patients. Figure 1 shows the trend of patients undergoing rehabilitation at our hospital, an overview of the number of patients, [21] and the utilization rate of COVID-19 beds [22] in Japan. The number of beds available to accept COVID-19 patients promptly ranged from 16,081 to a maximum of 39,792 nationwide [22].

**Table 1.**
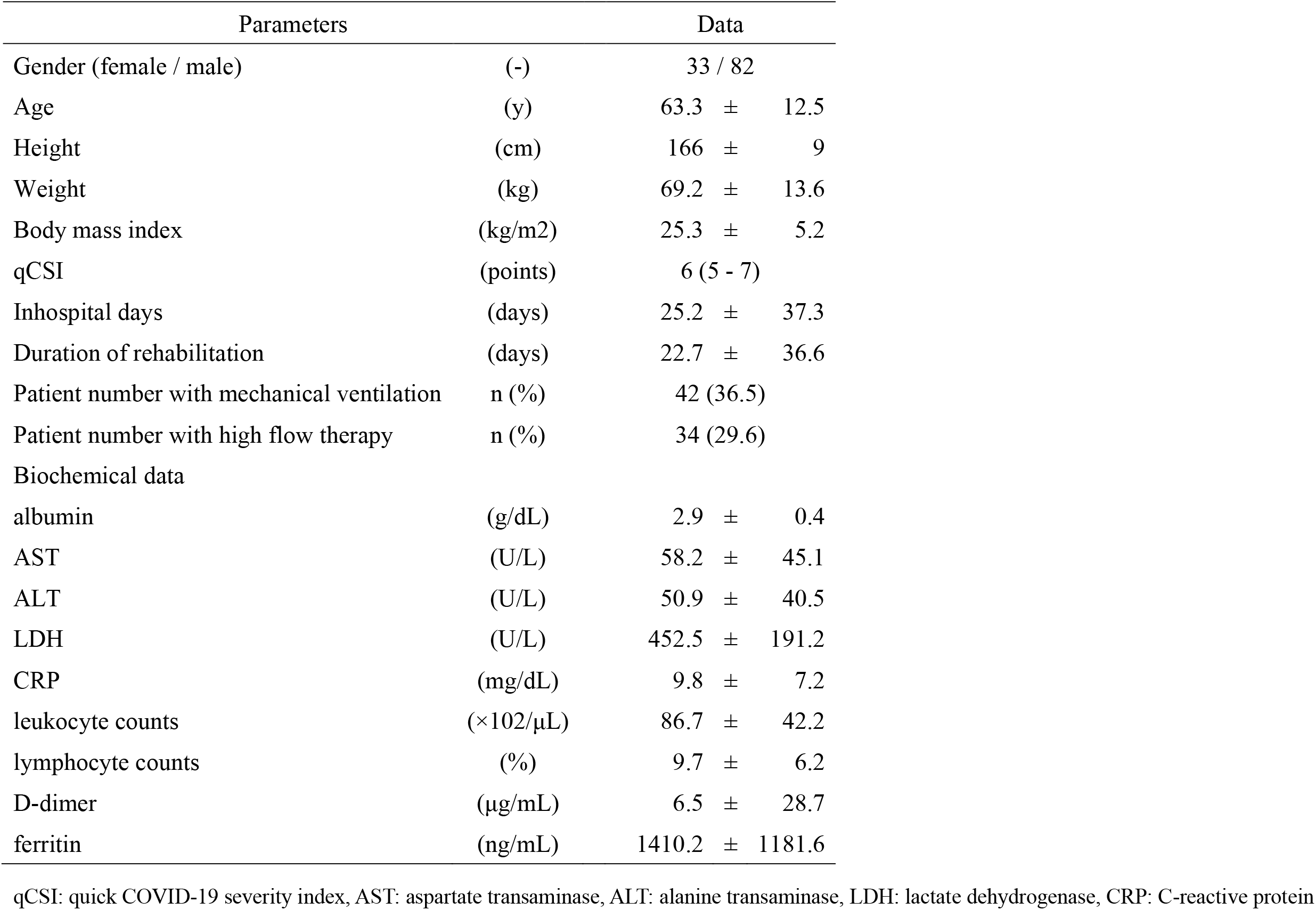
Patient characteristics of COVID-19

| Parameters |  | Data |  |  |
| --- | --- | --- | --- | --- |
| Gender (female / male) | (-) | 33 / 82 |  |  |
| Age | (y) | 63.3 | ± | 12.5 |
| Height | (cm) | 166 | ± | 9 |
| Weight | (kg) | 69.2 | ± | 13.6 |
| Body mass index | (kg/m2) | 25.3 | ± | 5.2 |
| qCSI | (points) | 6 (5 - 7) |  |  |
| Inhospital days | (days) | 25.2 | ± | 37.3 |
| Duration of rehabilitation | (days) | 22.7 | ± | 36.6 |
| Patient number with mechanical ventilation | n (%) | 42 (36.5) |  |  |
| Patient number with high flow therapy | n (%) | 34 (29.6) |  |  |
| Biochemical data |  |  |  |  |
| albumin | (g/dL) | 2.9 | ± | 0.4 |
| AST | (U/L) | 58.2 | ± | 45.1 |
| ALT | (U/L) | 50.9 | ± | 40.5 |
| LDH | (U/L) | 452.5 | ± | 191.2 |
| CRP | (mg/dL) | 9.8 | ± | 7.2 |
| leukocyte counts | (×102/μL) | 86.7 | ± | 42.2 |
| lymphocyte counts | (%) | 9.7 | ± | 6.2 |
| D-dimer | (μg/mL) | 6.5 | ± | 28.7 |
| ferritin | (ng/mL) | 1410.2 | ± | 1181.6 |

qCSI: quick COVID-19 severity index, AST: aspartate transaminase, ALT: alanine transaminase, LDH: lactate dehydrogenase, CRP: C-reactive protein

**Figure 1.**
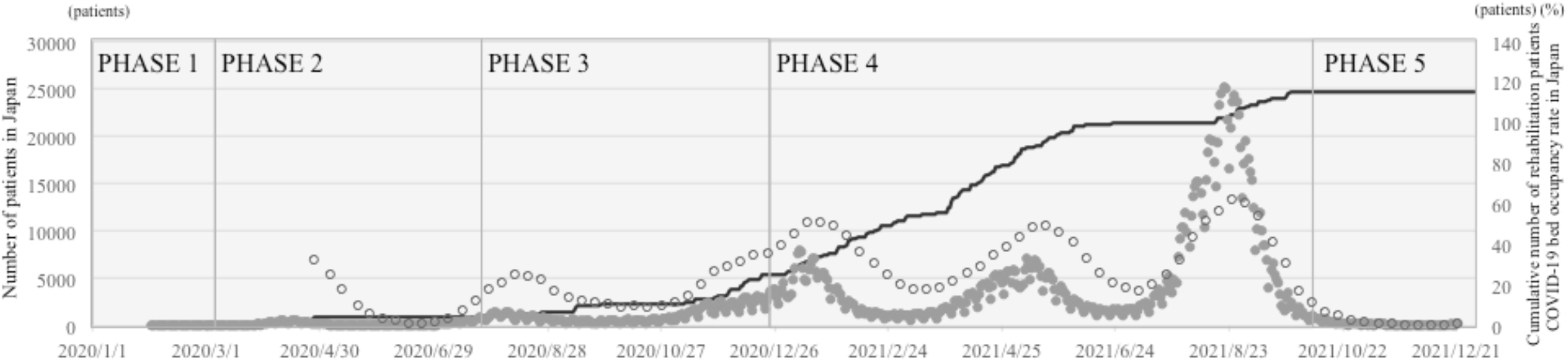
Cumulative number of rehabilitation patients in Hyogo College of Medicine and patient number of Japan. Gray dots represent patient number in Japan. (The graph was created from the public data on Our World Data website^21)^.) White dots represent COVID-19 bed occupancy rate in Japan. (The graphs were created from publicly available data on the Ministry of Health, Labour and Welfare website^22)^.) Black line represents cumulative number of rehabilitation patients with COVID-19 in our hospital.

The function of each phase, analysis of each aspect, and FRAM diagrams are shown below. FRAM functions are represented in Table 2 and FRAM diagrams are presented in Figures 2 and 3.

**Table 2.**
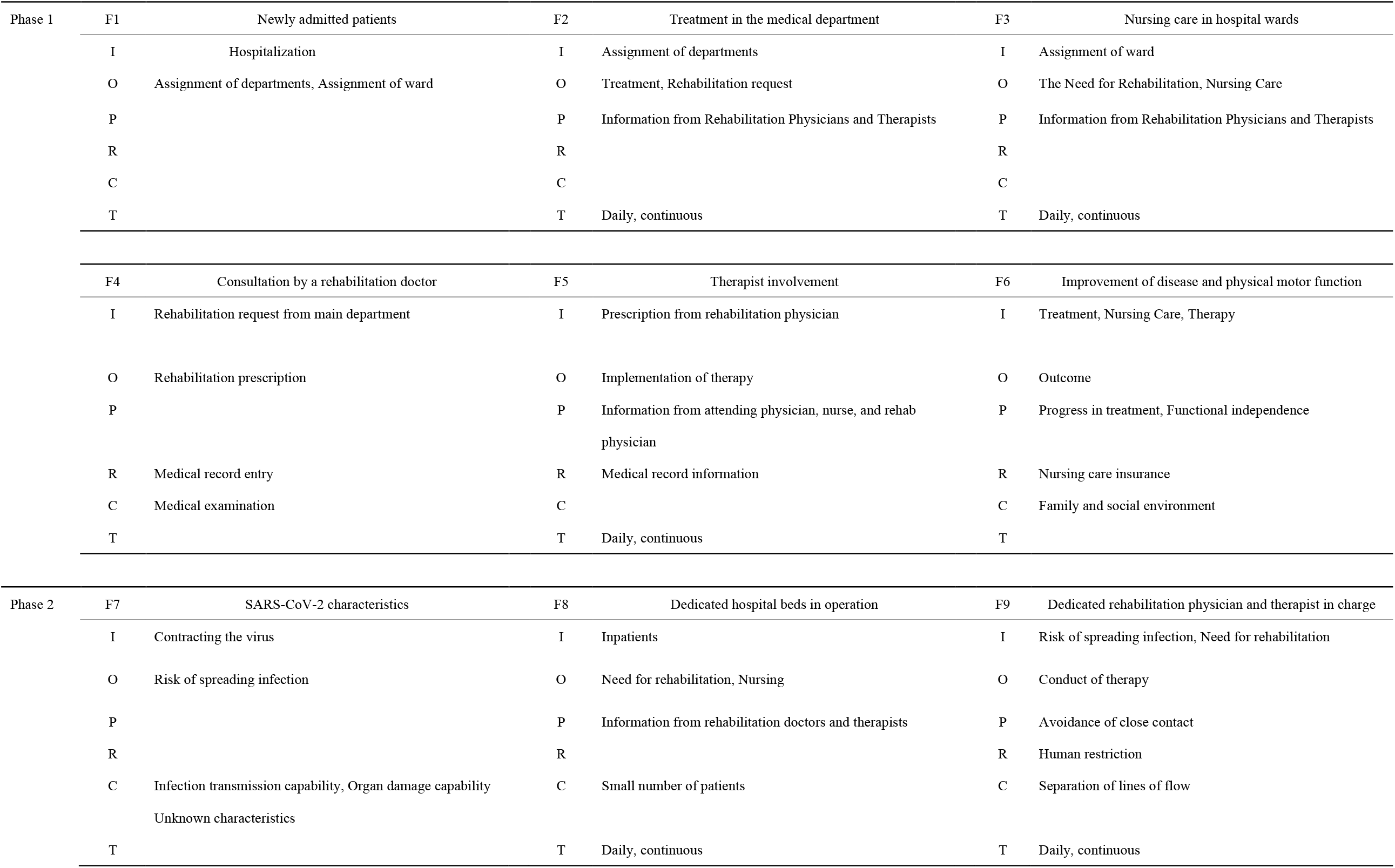
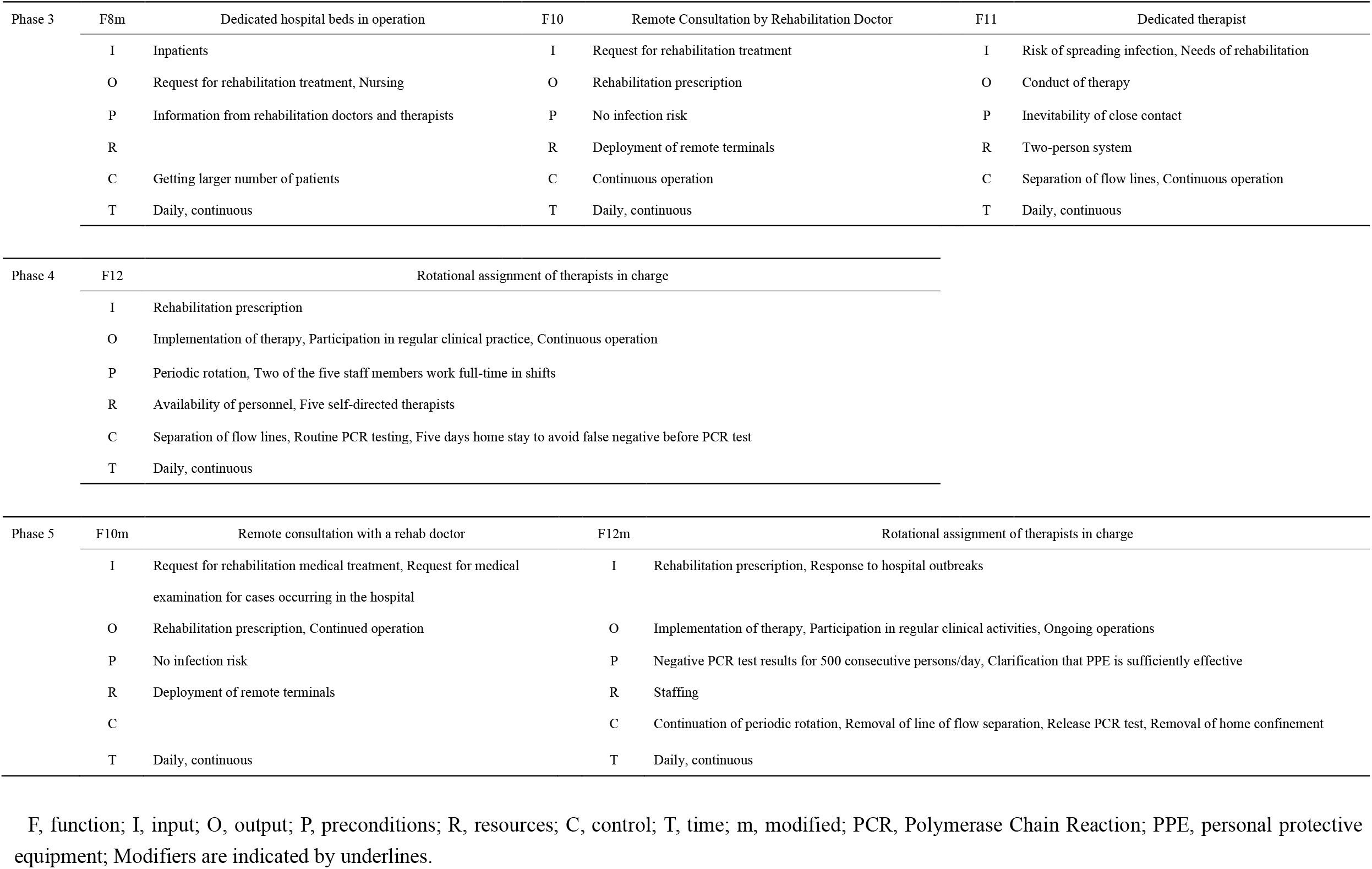
Description of functions and components

| Phase 1 | F1 | Newly admitted patients | F2 | Treatment in the medical department | F3 | Nursing care in hospital wards |
| --- | --- | --- | --- | --- | --- | --- |
|  | I | Hospitalization | I | Assignment of departments | I | Assignment of ward |
|  | O | Assignment of departments, Assignment of ward | O | Treatment, Rehabilitation request | O | The Need for Rehabilitation, Nursing Care |
|  | P |  | P | Information from Rehabilitation Physicians and Therapists | P | Information from Rehabilitation Physicians and Therapists |
|  | R |  | R |  | R |  |
|  | C |  | C |  | C |  |
|  | T |  | T | Daily, continuous | T | Daily, continuous |
|  | F4 | Consultation by a rehabilitation doctor | F5 | Therapist involvement | F6 | Improvement of disease and physical motor function |
|  | I | Rehabilitation request from main department | I | Prescription from rehabilitation physician | I | Treatment, Nursing Care, Therapy |
|  | O | Rehabilitation prescription | O | Implementation of therapy | O | Outcome |
|  | P |  | P | Information from attending physician, nurse, and rehab physician | P | Progress in treatment, Functional independence |
|  | R | Medical record entry | R | Medical record information | R | Nursing care insurance |
|  | C | Medical examination | C |  | C | Family and social environment |
|  | T |  | T | Daily, continuous | T |  |
| Phase 2 | F7 | SARS-CoV-2 characteristics | F8 | Dedicated hospital beds in operation | F9 | Dedicated rehabilitation physician and therapist in charge |
|  | I | Contracting the virus | I | Inpatients | I | Risk of spreading infection, Need for rehabilitation |
|  | O | Risk of spreading infection | O | Need for rehabilitation, Nursing | O | Conduct of therapy |
|  | P |  | P | Information from rehabilitation doctors and therapists | P | Avoidance of close contact |
|  | R |  | R |  | R | Human restriction |
|  | C | Infection transmission capability, Organ damage capability<br>Unknown characteristics | C | Small number of patients | C | Separation of lines of flow |
|  | T |  | T | Daily, continuous | T | Daily, continuous |
| Phase 3 | F8m | Dedicated hospital beds in operation | F10 | Remote Consultation by Rehabilitation Doctor | F11 | Dedicated therapist |
|  | I | Inpatients | I | Request for rehabilitation treatment | I | Risk of spreading infection, Needs of rehabilitation |
|  | O | Request for rehabilitation treatment, Nursing | O | Rehabilitation prescription | O | Conduct of therapy |
|  | P | Information from rehabilitation doctors and therapists | P | No infection risk | P | Inevitability of close contact |
|  | R |  | R | Deployment of remote terminals | R | Two-person system |
|  | C | Getting larger number of patients | C | Continuous operation | C | Separation of flow lines, Continuous operation |
|  | T | Daily, continuous | T | Daily, continuous | T | Daily, continuous |
| Phase 4 | F12 | Rotational assignment of therapists in charge |  |  |  |  |
|  | I | Rehabilitation prescription |  |  |  |  |
|  | O | Implementation of therapy, Participation in regular clinical practice, Continuous operation |  |  |  |  |
|  | P | Periodic rotation, Two of the five staff members work full-time in shifts |  |  |  |  |
|  | R | Availability of personnel, Five self-directed therapists |  |  |  |  |
|  | C | Separation of flow lines, Routine PCR testing, Five days home stay to avoid false negative before PCR test |  |  |  |  |
|  | T | Daily, continuous |  |  |  |  |
| Phase 5 | F10m | Remote consultation with a rehab doctor | F12m | Rotational assignment of therapists in charge |  |  |
|  | I | Request for rehabilitation medical treatment, Request for medical examination for cases occurring in the hospital | I | Rehabilitation prescription, Response to hospital outbreaks |  |  |
|  | O | Rehabilitation prescription, Continued operation | O | Implementation of therapy, Participation in regular clinical activities, Ongoing operations |  |  |
|  | P | No infection risk | P | Negative PCR test results for 500 consecutive persons/day, Clarification that PPE is sufficiently effective |  |  |
|  | R | Deployment of remote terminals | R | Staffing |  |  |
|  | C |  | C | Continuation of periodic rotation, Removal of line of flow separation, Release PCR test, Removal of home confinement |  |  |
|  | T | Daily, continuous | T | Daily, continuous |  |  |

F, function; I, input; O, output; P, preconditions; R, resources; C, control; T, time; m, modified; PCR, Polymerase Chain Reaction; PPE, personal protective equipment; Modifiers are indicated by underlines.

**Figure 2.**
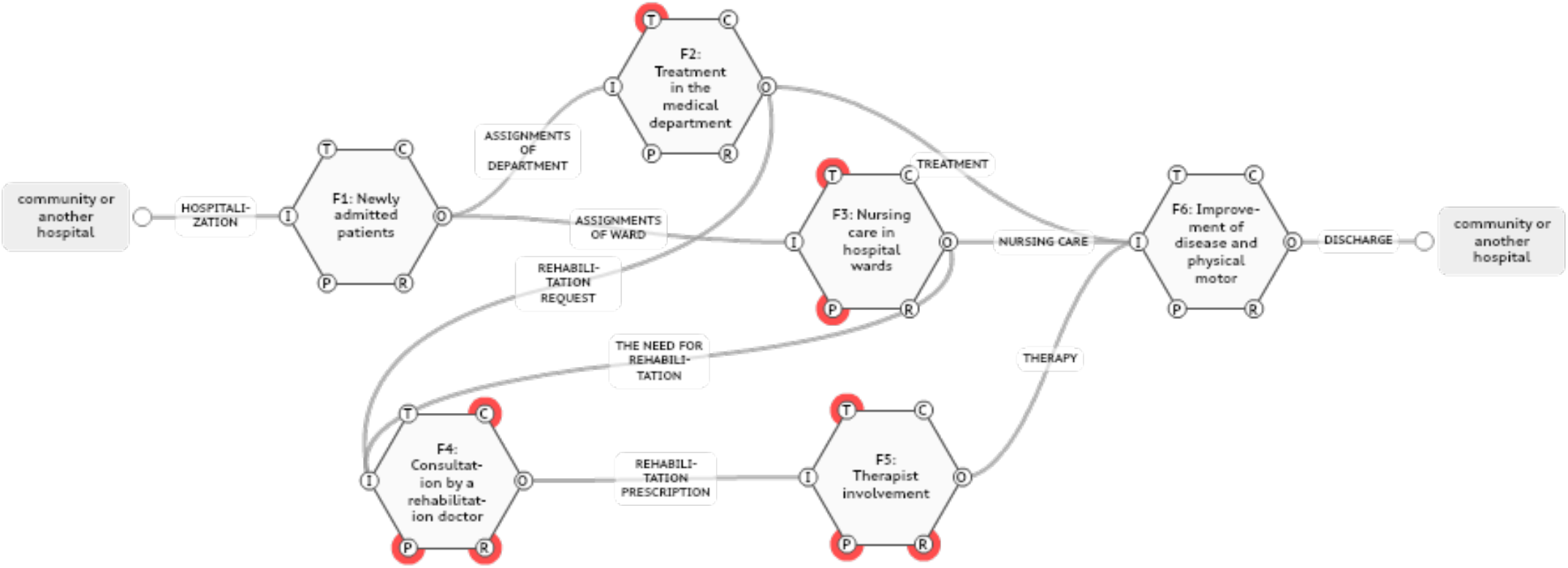
FRAM structure in PHASE 1 F; function, I; input, O; output, P; preconditions, R; resources, C; control, T; time, Red circles represent aspects that each function implements.

**Figure 3.**
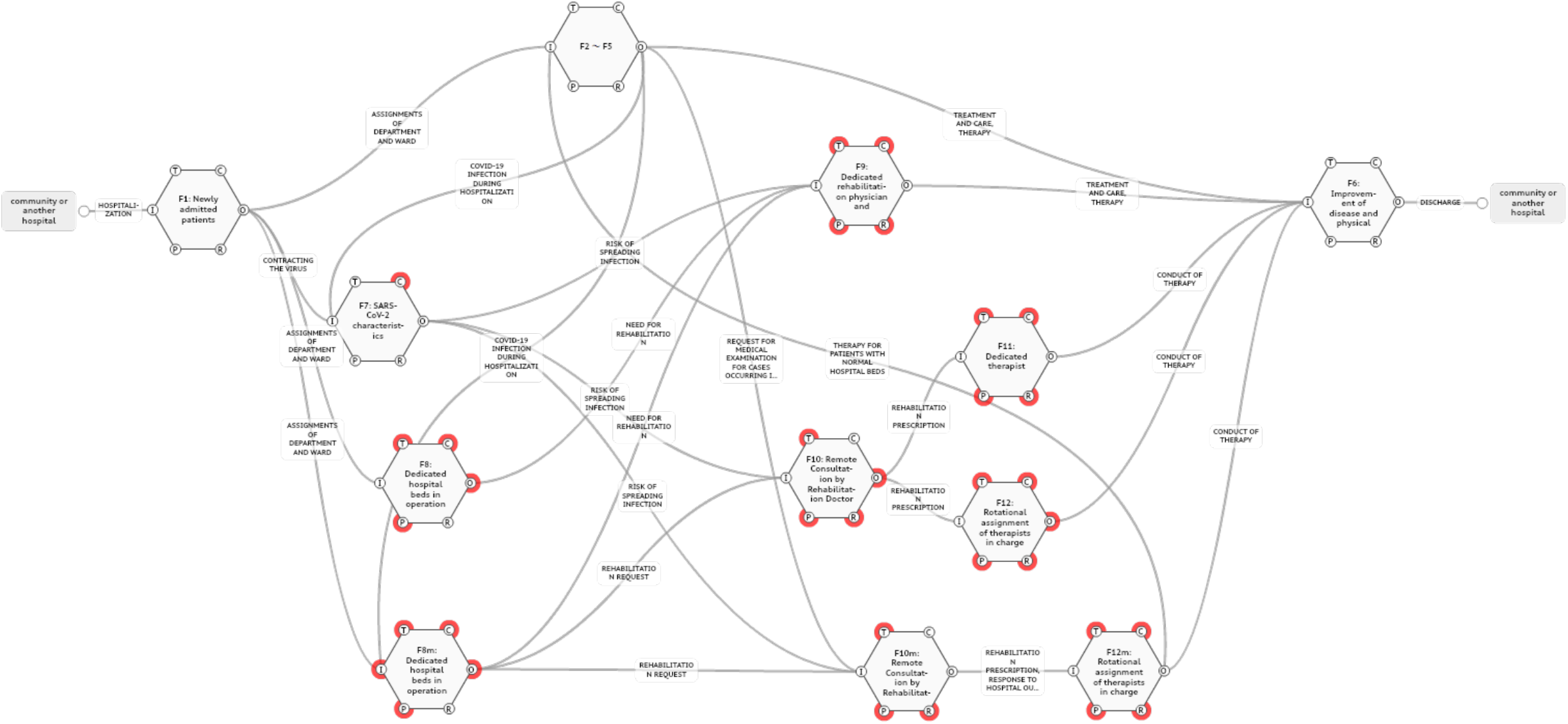
FRAM structure in Phase 2 – Phase 5 F; function, I; input, O; output, P; preconditions, R; resources, C; control, T; time, Red circles represent aspects that each function implements.

### Phase 1. Analysis of the provision system for medical rehabilitation in normal operations

When a patient is admitted to the hospital, he or she is assigned to a department and an inpatient ward, and the main department treats the main condition. In the rehabilitation of hospitalized patients, a rehabilitation prescription is issued after consultation by a rehabilitation physician in parallel with the treatment in the main department, and the therapist implements therapy based on the rehabilitation prescription.

### Phase 2. Analysis at 3 months after acceptance of COVID-19 cases

The hospital began accepting only patients with severe COVID-19. The greatest concern while providing rehabilitation therapy was that the therapists themselves could become infected and become the starting point for cross-infection and clusters in the hospital. Because of the high contact density of therapists with patients, the limited supply of PPE, and the small number of patients admitted with COVID-19 (maximum of 4), we decided to rehabilitate COVID-19 cases with the minimum number of therapists (one therapist).

### Phase 3. Analysis from 4 to 8 months

The number of hospitalized patients increased as the number of cases with COVID-19 in Japan gradually increased. During this period, four beds were always occupied, and the severity of illness gradually increased. As the number of patients increased, the number of COVID-19 beds was increased to six. In this phase, there was an increase in the burden associated with having only one therapist for each COVID-19 patient while maintaining a dedicated rehabilitation physician-therapist system. This increase in burden was due to cases that required a mechanical ventilator or were of a large physique and required much effort to reposition, as well as training required for mobilization intervention after sedation.

As a specific change in the functional resonance analysis, an increase in the number of hospital beds was added to the Resources in F8 (F8m; Fig. 3). F10 was added to F9 to allow rehabilitation physicians to see patients in an environment with no risk of infection, especially because of the limited number of staff.

The therapist’s response was to set F11 instead of F9 (Fig. 3). In F11, one therapist was added to the “Resources”, creating a two-person team. In this case, the report of the person in charge and the monitoring of bed turnover by the team manager functioned as “Control”.

### Phase 4. Analysis at Months from 9th to 21st

As a more permanent measure for patients with COVID-19, the hospital substantially increased the number of COVID-19 beds (up to 18 beds). Since the policy of accepting only severely ill patients was continued, two of the five dedicated therapists were rotated to provide continuous and safe rehabilitation intervention to the ten severely ill cases with COVID-19. Since the risk of transmission increased with the emergence of mutant strains and the combination of contact density and infectivity of therapists remained unclear, we established a system in which therapists were separated from other therapists from the time they arrived at work to the time they left work, and polymerase chain reaction (PCR) testing was performed routinely after a certain period of time (approximately 3 weeks) of patient care. After confirmation of negative results, the therapists concerned were to engage in normal work in general hospital beds. F12 replaced F11 in functional resonance analysis (Fig. 3).

### Phase 5. Consideration of challenges expected after the 22nd month (at present) and planning of countermeasures

At this point, in the 22nd month, we have identified actions that are expected to be needed and the measures that will be taken. The above system was continued for about 13 months, but PCR negative results continued for a total of 18 months (9 months × 2 therapists). Therefore, we concluded that the PPE used by the dedicated therapists on a daily basis was functioning well and routine PCR testing was discontinued. In addition, there is a need to establish a more permanent and flexible system to cope with the increased number of mild and moderate cases occurring in the hospital and the periodic or sudden replacement of dedicated therapists. In the functional resonance analysis, F12 was modified to F12m (Fig. 3).

## DISCUSSION

In this study, we used FRAM to analyze retrospectively the system used to provide safe and uninterrupted medical rehabilitation for patients with severe COVID-19 and to describe the problems of the system, as well as improvement measures derived from the FRAM analysis.

We used FRAM to identify problems and analyze system elements that should be continued and those that need to be modified or improved in order to establish a medical rehabilitation system for patients with COVID-19. To the best of our knowledge, this is the first report of a FRAM analysis of the system providing rehabilitation in a medical setting under the conditions of a SARS-CoV-2 outbreak.

FRAM is a systems analysis method [12], presented by Hollnagel E. in 2004, for clarifying and analyzing factors including rationality, validity, and “what works” by focusing on the interaction of the various functions that make up the system, rather than simply focusing on cause-and-effect relationships, abnormalities, and errors in the system. FRAM is comprehensive in that it considers a system from six aspects.

Healthcare systems are defined as complex adaptive systems [23]. A complex adaptive system is one that has more than a few variables or inputs, shows a nonlinear behavior, and interacts with its environment[23]. A health care system functions to achieve a goal while continuously adapting to the environment under constantly changing conditions [24]. FRAM is considered to represent an approach to nonlinear models, and we believe that it is useful for analysis of health care systems.

Figure 1 shows that the number of patients admitted to our hospital and the utilization rate of domestic hospital beds are linked to changes in the number of infected people in Japan. Table 1 shows that the population of 115 patients in this study required high-flow oxygen therapy or a mechanical ventilator in 76 cases (66%), indicating that the population included many moderately to severely ill patients and that, based on BMI, the population showed a larger physique compared with the standard body size in Japan [25]. Furthermore, Figure 1 shows that most of these patients were identified before October 2021, with few new cases for rehabilitation after November 2021. Vaccination is being administered on a global scale [26], and, in Japan, vaccination began in March 2021, with 73.4% of the population having completed the second vaccination by the end of December 2021 [27]. It is assumed that widespread vaccination has contributed to the reduction in the number of infected cases in Japan and the number of hospitalized patients in our hospital.

For each phase, the process of modifying the construction of the system providing medical rehabilitation for patients with COVID-19 was analyzed using the functional resonance method, and a functional table and FRAM diagram for each phase were created.

An analysis of the normal operating system shown in Phase 1 and the system under the COVID-19 shown in Phase 2 showed that one key factor is the potential infection risk of the virus prior to intervention by therapists. The FRAM diagram shows that the ability of the virus to transmit infection, preceding rehabilitation, has a significant impact.

In Phase 2, the characteristics of SARS-CoV-2 were defined as F7. Hollnagel defines “Function” as “the means that are necessary to achieve a goal “[28]. According to this definition, the “viral characteristics” of F7 are ineligible and may have to be shown as “preconditions” for F1. However, the characteristics of the current SARS-CoV-2 virus itself have changed, and the ease of infection and the properties of organ damage associated with infection are now variable. Therefore, in this report, we defined viral characteristics as a “Function “to show the two variables (infectivity and organ-damaging-properties) that characterize the virus as “Control”. However, it may be necessary to indicate each of these properties as an individual “function” because these properties themselves are variable, such as the ability to transmit infection, organ-damaging-properties, or other unknown characteristics.

In Phase 2, we incorporated the physician and therapist as a “Function” of F9. However, it may be more appropriate to treat these human factors as “Resources” in the system. It is well known that health care involves many professions [29]. The duties of each professional are strictly defined in accordance with the individual laws and regulations. On the other hand, this “definition” represents a specialized skill allowed only to each profession, and the function that each profession performs in the medical team. For this reason, in this study, we positioned professional occupations as “Functions” and analyzed each.

F10 in Phase 3 has remote consultation as a Function, and “Preconditions” in F10 is “no risk of infection”. This is a description of the situation associated with performing the duties of a medical examination, and does not include general risks throughout daily life. F11 can be regarded as a modified F9, but in this analysis, it is positioned as a different function from F9 because the type of work involved has changed.

At F11, two therapists began rehabilitation intervention in patients with COVID-19, and, at F12 in Phase 4, five more therapists were assigned to patients with COVID-19, with two of these therapists shifted to take charge of COVID-19 beds. The main features of F12 are the PCR test given to the therapist in charge and the 5-day home waiting period before the test to avoid false negatives. This routine PCR test was performed as a means of avoiding the therapist becoming the source of the cluster when the risk of infection to the therapist was unknown.

In Phase 6, the F12m system was established as a more permanent system, and regular PCR testing of dedicated therapists was halted because no dedicated therapists were affected by COVID-19 during the 540 person-days of intervention in COVID-19 beds. Thus, although we considered the negative PCR results of 540 persons · days and 27 tests of dedicated personnel as the basis for the termination of periodic PCR testing, it is difficult to interpret whether the termination of periodic testing was reasonable. In a previous study, the risk avoidance rate of COVID-19 infection with PPE was reported to be >95% (26 negative results in 27 PCR tests) [30], while our test results showed 27 negative results in 27 PCRs (100%). These results indicate that daily PPE wearing by each therapist functions adequately, and our response was appropriate.

F12m was analyzed as a prospective forecast and future measures were discussed. A prospective study using FRAM [31] reported on the establishment of a wide-area medical system for COVID-19, and described its usefulness for predictive scenario analysis. F12m was developed based on the analysis of possible future disturbances, but it may not be able to cope with disturbances that occur suddenly and without warning.

There are some limitations to this study. The setting and description of each “Function” has a high degree of freedom, and the reproducibility and universality of the model may be poor. In addition, since the scope of each function acting on the entire system is infinitely broad and can be infinitely subdivided, it is essential to examine the validity of the breadth and fineness of the descriptions in this report. Although we have attempted to formulate some prospective functions, the analysis of disturbances that occur suddenly and without warning must be performed retrospectively. The usefulness of FRAM from a predictive perspective is an important issue for the future.

In this study, we conducted FRAM system analysis of the process of establishing a safe and uninterrupted system to provide rehabilitation for COVID-19 patients and attempted to develop a strategy to deal with foreseeable problems. The presentation of “Functions” and the analysis of the elements of each “Function” are useful for constructing a system can be modified from time to time, and may be applied to the operation of different systems in different situations. From our results, we conclude that the analysis of our medical rehabilitation provision system by FRAM allows the prediction of necessary measures in the future and contributes to the establishment of a safe and uninterrupted system for providing rehabilitation.

## Data Availability

All data produced in the present study are available upon reasonable request to the authors.

## SQUIRE 2.0 CHECKLIST

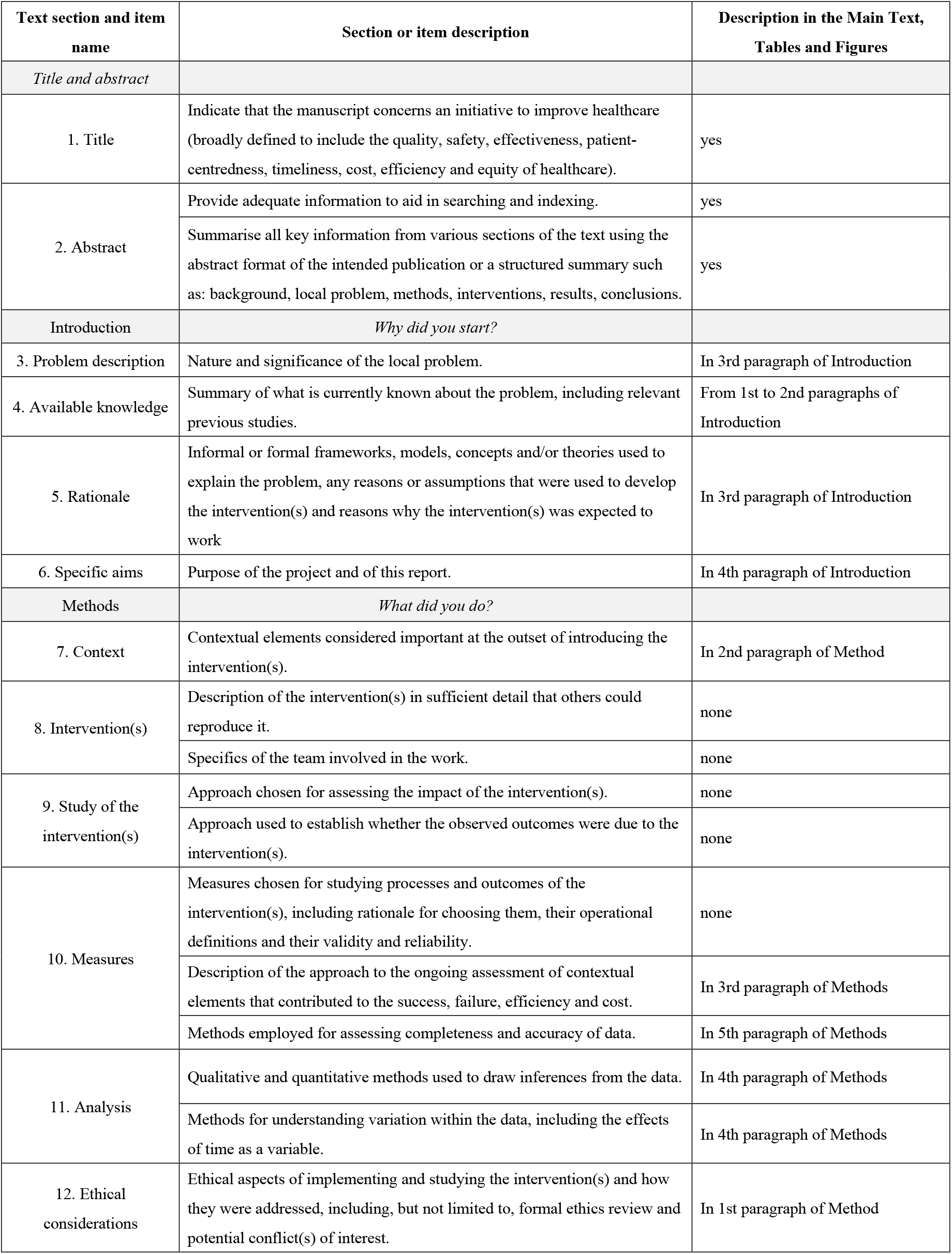
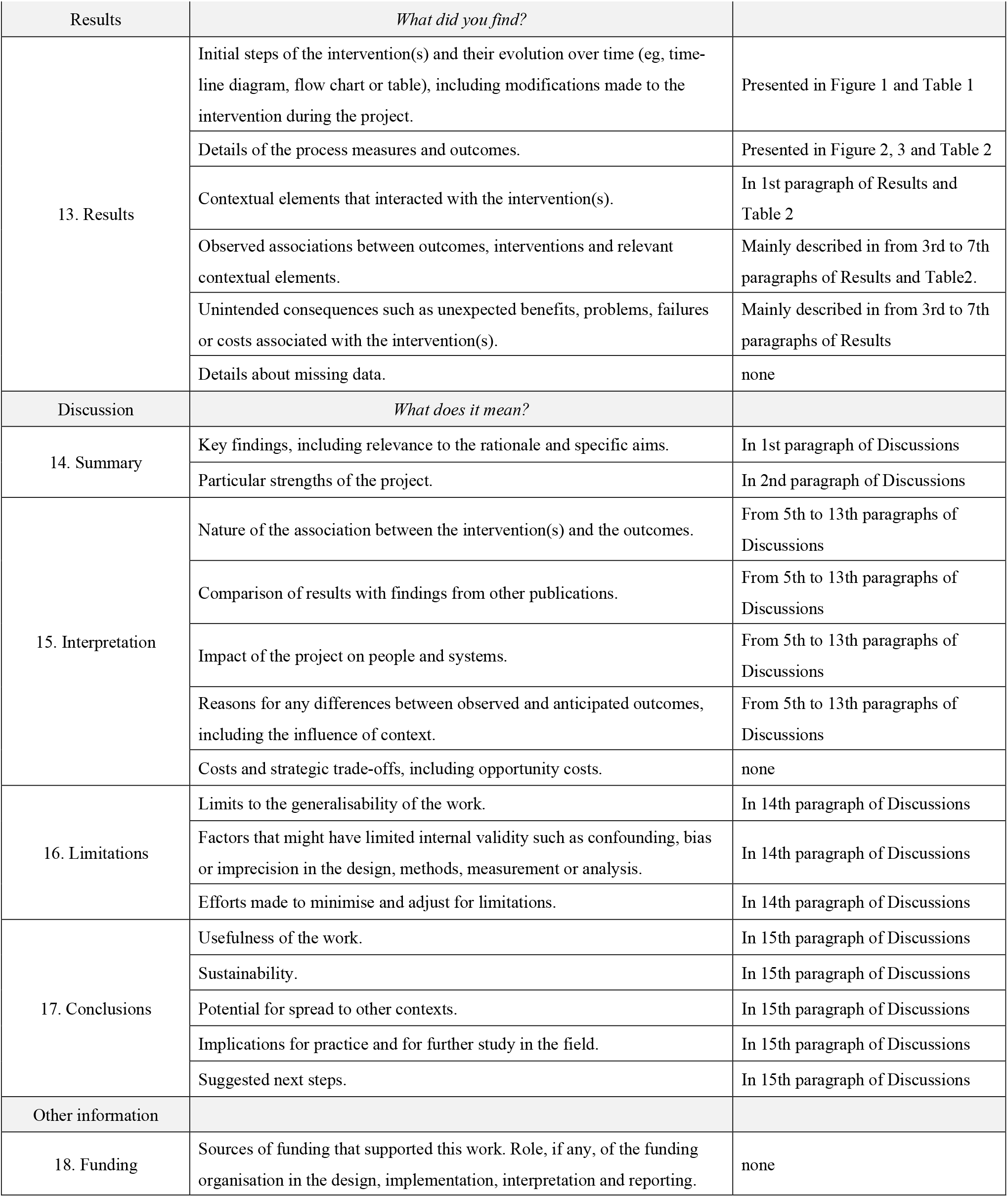

